# Large-scale deployment of SIT-based technology in a Brazilian city prevented Dengue outbreak

**DOI:** 10.1101/2022.09.19.22279924

**Authors:** Lisiane de Castro Poncio, Filipe Apolinário dos Anjos, Deborah A. de Oliveira, Aline Rosa, Bianca Piraccini Silva, Débora Rebechi, Diego Alan da Costa Franciscato, Cláudio de Souza, Uilson Paiva, Marilu Mazurechen, Rafael de Araújo Ribeiro, Priscila Basile, Erikon Leandro Rezende, Nitzan Paldi

## Abstract

**Background:** Dengue is a global problem that seems to be worsening, as hyper-urbanization associated with climate change has led to a significant increase in the abundance and geographical spread of its principal vector, the Ae*des aegypti* mosquito. The current available solutions, including vaccines and traditional vector-control methods, have not been able to stop the spread of dengue which shows the urgent need to implement alternative technologies as practical solutions. We recently presented ‘Natural Vector Control’ (NVC), a new Sterile Insect Technology-based method that uses massive releases of sterile male mosquitoes produced from the combined treatment with dsRNA and thiotepa. In a previous pilot trial, two intervention periods over two epidemiological seasons were carried out, in which the control and treated areas were alternated between the epidemiological seasons, and we demonstrated the efficacy and safety of the method in suppressing the *Ae. aegypti* vector population and in blocking the occurrence of an outbreak of dengue in the treated areas. Here, we expand the use of the “Natural Vector Control” program in a large-scale 2-year period intervention carried out in an entire city located in southern Brazil.

**Methods:** Sterile male mosquitoes were produced from locally sourced *Ae. aegypti* mosquitoes by using a treatment that includes double-stranded RNA and thiotepa. Weekly massive releases of sterile male mosquitoes were performed in predefined areas of Ortigueira from December 2020 to July 2022. Mosquito monitoring was performed by using ovitraps during the entire period of intervention. Dengue incidence data in Ortigueira and neighboring cities was obtained from the Brazilian National Disease Surveillance system.

**Results:** During the two epidemiological seasons, the intervention in Ortigueira resulted in up to 98.7% suppression of live progeny of field *Ae. aegypti* mosquitoes recorded over time. More importantly, the program protected Ortigueira from a dengue outbreak that occurred in the neighboring cities: the dengue incidence in Ortigueira was 97% lower compared to 4 control cities.

**Conclusions:** The Natural Vector Control method has again been shown to be a safe and efficient way to suppress *Ae. aegypti* field populations and prevent the occurrence of a dengue outbreak. Importantly, it has been shown to be applicable for large-scale, real-life conditions.

## Background

Dengue is a mosquito-borne viral disease that represents a massive and growing problem worldwide. Globally, its main vectors are mosquitoes of the specie *Ae. aegypti*. This mosquito is also the principal global vector of chikungunya, yellow fever and Zika (1). Historically, the incidence of arboviruses, especially dengue, was concentrated throughout the tropics (1). However, hyper-urbanization and resulting climate change from anthropological activities has resulted in the expansion of this mosquito’s population into regions that until recently were not infested (2,3). Subsequently, dengue and other *Ae. aegypti* transmitted arboviruses are becoming a concern to these regions (2–4).

Along with the global increase in the incidence of dengue, cases of severe dengue have also proportionally increased, which has a drastic impact on the morbidity and mortality of affected populations and important social and economic implications (2,5). To date, interventions such as vaccination have failed to control the spread of these arboviruses (other than yellow fever), and it is clear that effective and safe vector elimination is the key to reaching the WHO goals for dengue, i.e. reducing its global incidence by 75% until 2027 (6,7).

Sterile Insect Technology (SIT) to achieve population suppression, has emerged in the last decade as a highly effective and ecologically viable method to constrain the spread of disease-carrying mosquitoes. This technique, in its various manifestations, is based on the massive release of sterile male mosquitoes in the intervention area, which will mate with wild females and lead to a progressive reduction of mosquitoes in subsequent generations (8). The techniques used to date to reduce mosquito populations include the sterilization of mosquitoes by irradiation (8), the use of genetically modified mosquitoes carrying a dominant lethal gene (9), and incompatible insect technology, which utilizes mosquitoes infected with *Wolbachia* bacteria (10–15).

We previously provided direct evidence of prevention of dengue using a technique called Natural Vector Control (NVC) (16). We demonstrated that reversal of treatment and control areas in two neighborhoods in a Brazilian city, over two epidemiological seasons, brought >90% reduction of dengue cases in the treatment areas in the midst of a dengue epidemic in both seasons (16). Herein we present the results over two seasons of an intervention with large-scale deployment of NVC in Ortigueira, a city located in the southern region of Brazil. In early 2020, the city, as well as much of the state of Parana, experienced a major dengue outbreak (17). The massive releases of NVC sterile male mosquitoes started in December 2020 and lasted until July 2022. This intervention drastically suppressed the infestation of *Ae. aegypti* and protected the city from another dengue outbreak that affected the entire State of Parana and the region around Ortigueira in the first seven months of -2022. Our results provide further support for NVC intervention as an effective method to prevent dengue that can be applied in virtually any city.

## Methods

### Production of NVC Sterile Male Mosquitoes

The *Ae. aegypti* colony was established from eggs collected in the northern region of Paraná, Brazil (2017), using ovitraps (18). Mosquitoes were reared essentially as described previously (16).

The NVC sterile male mosquitos’ large-scale production process was performed essentially as described in de Castro Poncio, 2021 (16). All the batches of NVC sterile male mosquitoes underwent a quality control to detect the potential presence of contaminating females and verify the sterility of the males.

### NVC sterile male mosquito releases and monitoring field Ae. aegypti mosquitoes

Ortigueira is a city in the northern region of Parana state, southern Brazil, with a population of 21 thousand inhabitants (source: 2010 Brazilian census). It has a subtropical climate, with an average temperature varying from 19.1 – 20 °C over the year. Releases of NVC mosquitoes were performed on a weekly basis from November 2020 to July 2022. NVC male mosquitoes were packed in plastic containers and manually released from a car driving along streets of the city, as previously described (16). The number of NVC male mosquitoes released in each microregion of the city was defined based on the size of each area and monitoring data of eggs and larvae collected in the areas in the previous week.

Monitoring of local *Ae. aegypti* abundance was performed by weekly installation of 159 ovitraps in predefined houses or in the peridomiciliary area of the residences in Ortigueira (Supplementary Figure 1) (16,19,20). Eggs from each ovitrap were counted and hatched for a 48-hour period. The mean number of larvae/ovitrap that hatched per in this period were considered viable progeny. The mosquito monitoring started one month before the beginning of NVC releases and continued until one month after the NVC intervention period.

Estimation of mosquito field population suppression was performed as previously described (16,21). Londrina city was used as a control city for *Ae. aegypti* monitoring by using ovitraps, similarly as described for Ortigueira. Briefly, monthly moving averages relative to the same period of the control area were calculated according to the equation M = (T_a_/C_a_)/(T_b_/C_b_) – 1, where M is the viable progeny change, T_a_ is mean larvae per point in the intervention area (Ortigueira) after release, C_a_ is mean larvae per point in the control area (Londrina) after release, T_b_ is mean larvae per point in the treated area before release, and C_b_ is mean larvae per point in the control area before release. This was done by comparing monthly data against baseline data obtained across the 2 months prior to the beginning of releases. The corresponding 95% confidence intervals (CIs) were calculated by a 10,000-loop bootstrap (21) for the entire period of releases and for each period of 4 weeks, to follow the effect along the project and considering the similar effect in coming weeks. The CIs were calculated using R version 4.2.1(R version 4.2.1 - 2022-06-23 ucrt - Copyright © 2022 The R Foundation for Statistical Computing).

### Epidemiological data

Epidemiological data regarding dengue cases in Ortigueira were obtained from the Brazilian National Disease Surveillance system (SINAN). Data on dengue is available on SINAN since 2000. Suspected cases of dengue are reported according to a case definition of fever plus at least two of the following additional symptoms: malaise, headache, myalgia, nausea, vomiting, cutaneous rash and arthralgia (15). Most suspected cases were tested by ELISA (IgM and IgG serology). PCR testing is performed only for severe and fatal cases, pregnant women and young children (22).

To assess the impact of the NVC intervention on the incidence of dengue in Ortigueira, epidemiological data on dengue in the cities neighboring Ortigueira (Marilância do Sul, Tamarana, Telêmaco Borba, Mauá da Serra, Apucarana, Imbaú, Londrina, Grandes Rios and Tibagi) were also collected. Dengue incidence in indicated periods was calculated as (number of dengue cases/ number of inhabitants) × 100,000. The number of inhabitants in each city was based on demographic data from Brazilian Institute of Geography and Statistics (IBGE). To evaluate the protective effect of NVC intervention on dengue incidence, the rate ratio (RR) was calculated (RR = I_DT_ / I_DC_). Where I_DT_ is the incidence of dengue cases in the treated area (Ortigueira) and I_DC_ is the dengue incidence of control cities during the period of intervention. Values of RR < 1 indicate that the NVC treatment is protective against dengue; 95% CIs were calculated using R software.

Estimation of dengue suppression after NVC intervention was performed using the 10,000 Loop Bootstrap simulation using 3-month period of comparison, starting in January 2020, one year before the intervention (16,21). Telêmaco Borba, Grandes Rios, Imbaú and Tibagi were used as control cities for dengue incidence in the same period of NVC intervention performed in Ortigueira. Monthly moving averages relative to the same period at control cities were calculated according to the equation M = (T_a_/C_a_)/(T_b_/C_b_) – 1, where M is the dengue incidence change, T_a_ is dengue incidence in Ortigueira after NVC releases, C_a_ is the incidence in the control cities after release, T_b_ is dengue incidence in Ortigueira before NVC releases, and C_b_ is dengue incidence in the control cities before NVC period releases. This was done by comparing 3-month data intervals against baseline data obtained across the 12 months prior to the beginning of releases. The corresponding 95% confidence intervals (CIs) were calculated by a 10,000-loop bootstrap (21) for the entire period of releases and for each period of 3 months, to follow the effect along the project and considering the similar effect in coming months. The CIs were calculated using R version 4.2.1(R version 4.2.1 - 2022-06-23 ucrt - Copyright © 2022 The R Foundation for Statistical Computing).

### Ethics and trial registration

An Environmental License from the Environmental Institute of Paraná was obtained (license number 36127) to perform the NVC intervention in Ortigueira. In addition, consent to perform the intervention was given by the Sanitary and Epidemiological Surveillance service the Health Secretariat and the Ortigueira City Hall (consent letter 20201109_101236 and transport authorization 197/2020).

## Results

### Early releases and 2 years of sequential intervention are required to successfully suppress local Ae. aegypti mosquito population

To demonstrate the feasibility of implementing the NVC in an entire city and the impact of this intervention on the prevention of dengue, a disease that has great social, economic, and public health implications, the city of Ortigueira, in Paraná, was chosen to receive a 2-year intervention plan. A total of 59 million sterile male mosquitoes (NVC) were released during the period from November 2020 to July 2022, in 200 points of the city, and the monitoring of mosquito infestation was carried out with the placement of ovitraps in 159 points throughout the city (Supplementary material 1). The intervention in Ortigueira resulted in a reduction in viable progeny of *Ae. aegypti* from the 18^th^ week of intervention (Figure 1A). From then on, the viable progeny of mosquitoes drops to close to zero and remains low throughout the second year of intervention.

**Figure 1.**
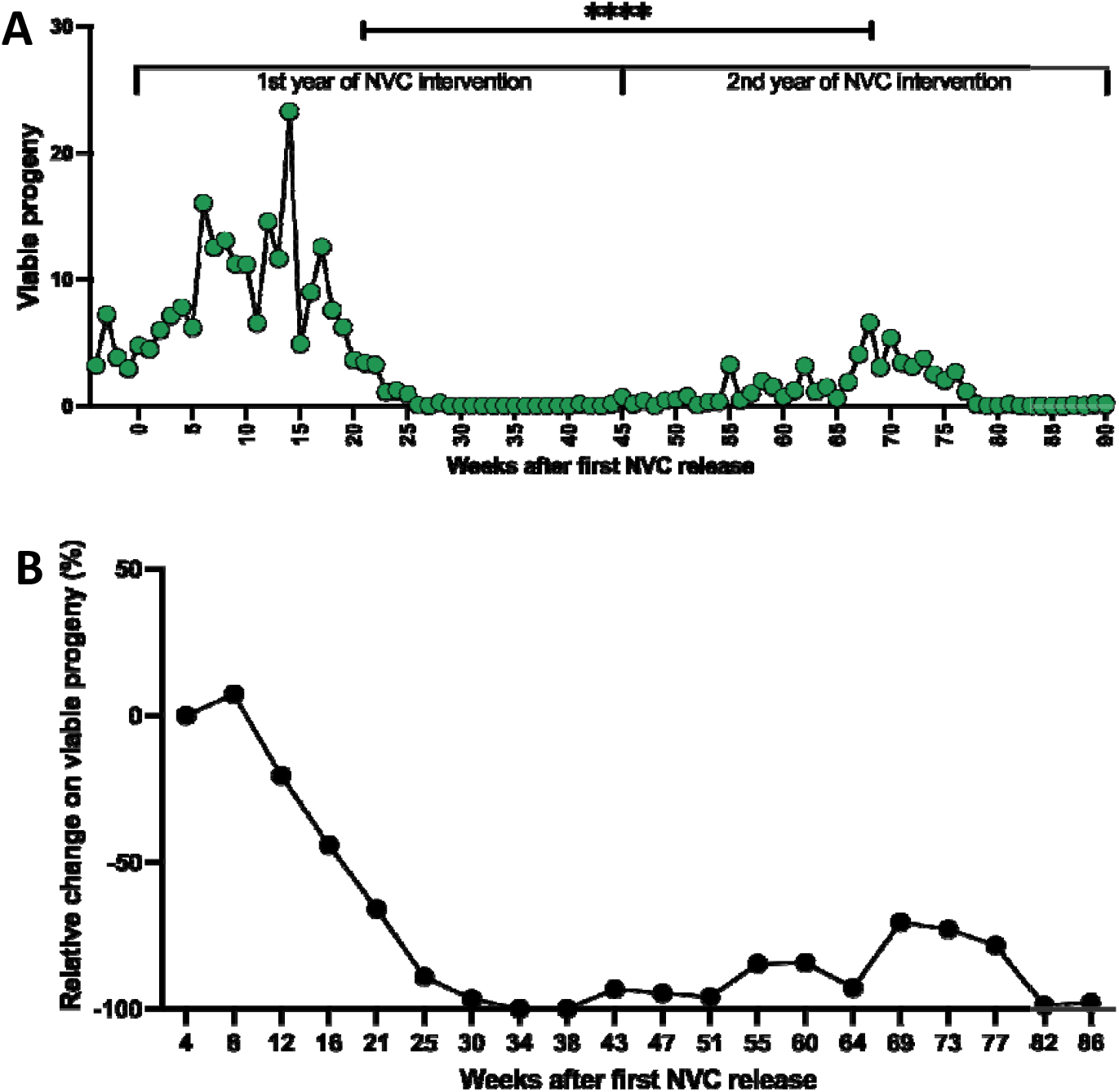
Suppression of field *Ae. aegypti* mosquito population in Ortigueira during 1^st^ and 2^nd^ years of NVC intervention. **A**. Releases of NVC sterile male mosquitoes occurred weekly between December 2020 and July 2022 (Weeks 0 to 90). The city was monitored with egg collection by using ovitraps during the entire period of NVC intervention. The collected eggs were transferred to the laboratory, where they were hatched. The mean number of larvae hatched from eggs of each ovitrap (159 points) over the 90 weeks was defined as viable progeny. Statistical analysis: Paired t test provided a P value of <0.0001) for the difference between 1^st^ and 2^nd^ year of NVC intervention. B. Suppression of *Ae. aegypti* wild population in Ortigueira during NVC program. Monthly moving averages showing percentage change in *Ae. aegypti* abundance in Ortigueira, measured by mean number of larvae per trap relative to control area (Londrina). In weeks 30-34, there was a 98.7% (95% CI, 98.3% to 100%) reduction in numbers of mosquitoes compared to 4.

To quantify the reduction in viable progeny of *Ae aegypti* in Ortigueira during the NVC intervention period, we performed the measurement of monthly moving averages of viable larval sampling hatched from eggs collected during field surveillances in both Ortigueira (intervention area) and Londrina (control area), a city near Ortigueira, as previously described (16,21). Over the 2 seasons of intervention, NVC dramatically reduced the *Ae. aegypti* mosquito field population in Ortigueira, reaching up to 98.7 % (CI – 98.3 % to – 100%) reduction in the number of viable larvae during the entire period of intervention (Figure 1B). Finally, a linear regression analysis showed a significant correlation between the release of NVC mosquitoes and the reduction in viable progeny (Supplementary material 2).

### Four neighboring cities present similar historical dengue incidence pattern over the past 20 years

To demonstrate that the NVC intervention actually prevented the occurrence of dengue in Ortigueira, we evaluated the incidence of dengue in Ortigueira throughout the intervention period, compared to the occurrence of dengue in other cities in the region. Ortigueira is surrounded by several other cities that have sociodemographic and climatic similarities (Supplementary material 3). The choice of cities to be used as controls was based mainly on the historical similarity of the occurrence of dengue outbreaks/or epidemics between the years 2000 and mid-2020, a period prior to the NVC intervention (15). We did not consider the history of other arboviruses (Zika and Chikungunya) as the incidence of these diseases in the region has been very low since these diseases began to be reported in Brazil (Supplementary Material 4). As can be seen in Figure 2, among the neighboring cities evaluated, Marilândia do Sul, Mauá da Serra, Londrina, Tamarana and Apucarana presented different dengue incidence peaks compared with the dengue peaks occurred in Ortigueira in the same period. On the other hand, the cities Grandes Rios, Telêmaco Borba, Imbaú and Tibagi showed similar patterns of dengue incidence over the last 2 decades when compared to Ortigueira, and therefore were chosen to serve as controls of dengue occurrence during and after the NVC intervention period.

**Figure 2.**
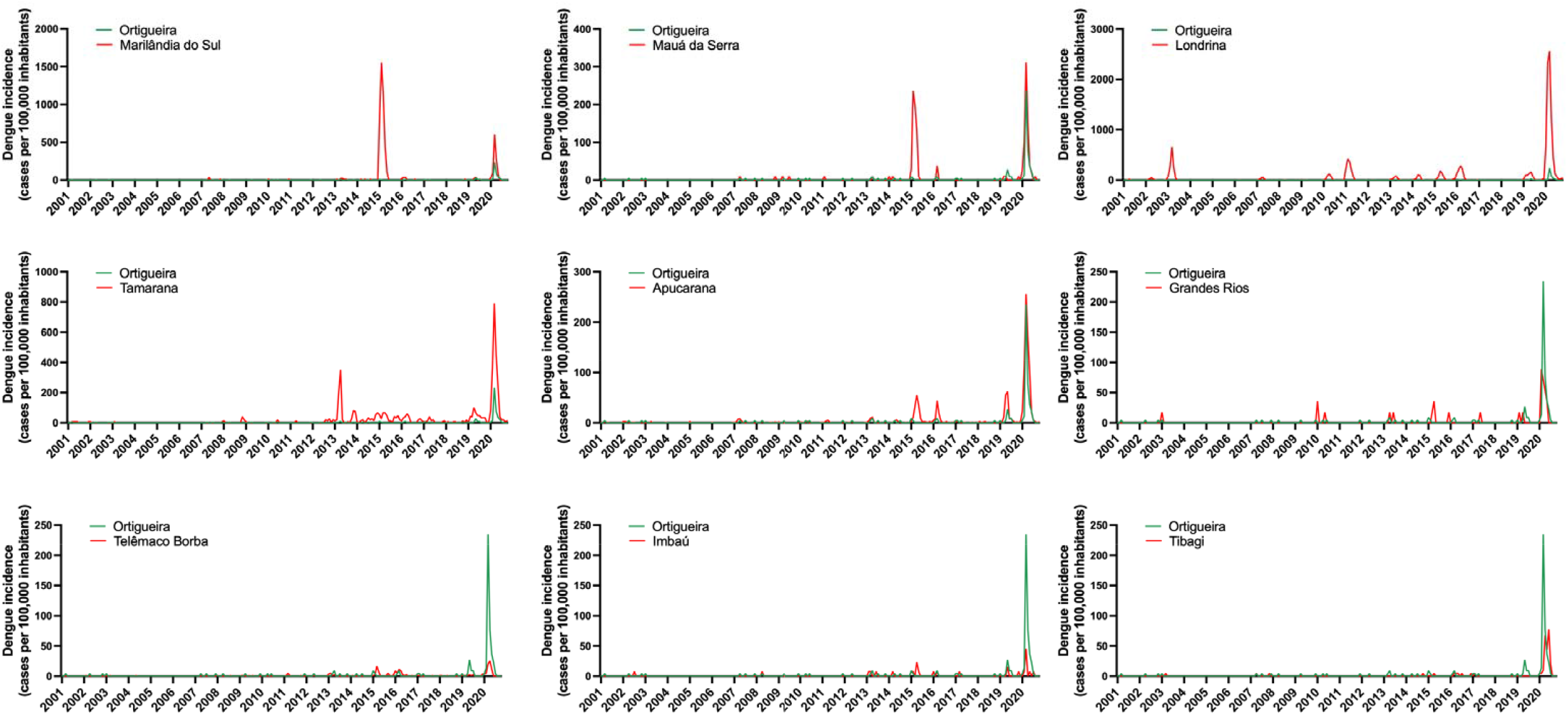
Historical dengue incidence in Ortigueira vs neighboring cities. Each graph shows dengue incidence from 2001 to 2020 (displayed as cases per 100,000 inhabitants) in Ortigueira (green line) compared with neighboring cities (red lines), as indicated (Marilândia do Sul, Mauá da Serra, Londrina, Tamarana, Apucarana, Grandes Rios, Telêmaco Borba, Imbaú, and Tibagi). Data source: Brazilian National Disease Surveillance System (SINAN).

### NVC intervention prevented an outbreak of dengue in Ortigueira

We had previously demonstrated the effectiveness of the NVC intervention in containing dengue outbreaks, by performing the reversion of the areas treated with the NVC and the control areas in 2 different epidemiological seasons, resulting in a substantial reduction in the dengue incidence in the treated areas (16). Herein we expanded the NVC intervention to an entire city. As we can see in Figure 3A, which shows the dengue incidence from Jan 2020 to July 2022, the first peak of incidence in the cities of the region (including Ortigueira) occurred in March 2020, before the NVC intervention period. The second and more important peak of dengue incidence occurred in May 2022. This period includes the NVC intervention performed in Ortigueira. The other cities nearby did not receive NVC intervention. with the incidence of dengue in the neighboring cities ranged from 120 cases per 100,000 inhabitants (Telêmaco Borba) to 3090 cases per 100,000 inhabitants (Tibagi). In contrast, Ortigueira, displayed a much lower dengue incidence (20 cases per 100,000 inhabitants) in the same period. There seems to be evidence that at least 3 of the 5 cases of dengue that occurred were contracted elsewhere (supplemental XX). Therefore, the incidence of dengue in Ortigueira was between 6-150 times lower in Ortigueira, compared to neighboring cities.

**Figure 3.**
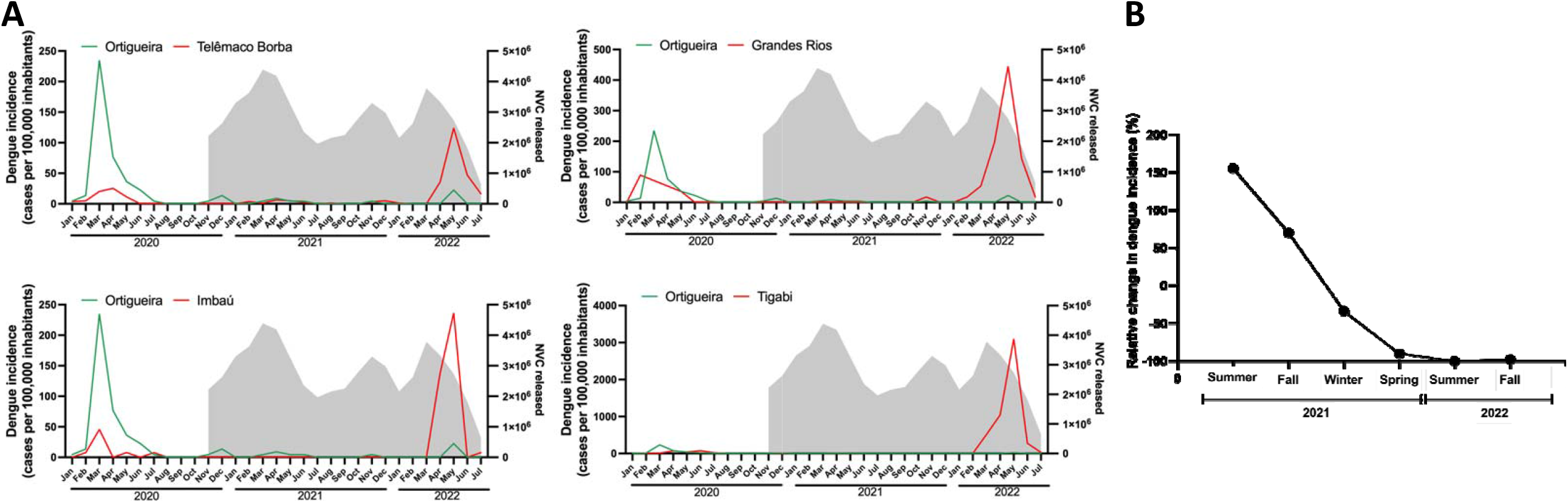
Dengue incidence in NVC intervention area (Ortigueira) and control neighboring cities from Jan 2020 to July2022. **A**. Dengue incidence in Ortigueira and the indicated control cities Telemaco Borba, Grandes Rios, Imbau and Tibagi is shown in the left axis, expressed as the number of dengue cases per 100,000 inhabitants. The grey shadows in each graph indicate the number of NVC sterile male mosquitoes monthly released in Ortigueira during the NVC period of intervention. The incidence of dengue in Ortigueira in May 2022 was 20 cases per 100,000 inhabitants (95% CI, 10-20), while the dengue incidence in control cities was 120 cases (95% CI, 100-150) in Telêmaco Borba; 440 cases (95% CI, 300-660) in Grandes Rios; 240 cases (95% CI, 170-340) in Imbaú; and 3090 cases (95% CI, 2870-3340) in Tibagi. **B**. Monthly moving averages of dengue incidence in Ortigueira and control cities. A 10,000 bootstrap simulation was performed to show the percentage change in dengue incidence in Ortigueira, using the dengue incidence of the four neighboring cities as controls. In the Spring of 2021, there was 89,85% reduction in the dengue incidence in Ortigueira compared to Winter 2021 (95% CI, 79.7% to 100%). In the Summer of 2022, the reduction on dengue incidence reached 100% compared to the previous season (Spring 2021) and kept low also in the Fall 2022.

The protective effect of NVC was confirmed calculating the dengue ratio between areas. The RR showed a reduction of 89.1 % in the incidence of dengue in Ortigueira compared to the other four control cities (CI −95.5 % to – 82.3 %) during the two years of intervention.

Finally, to assess whether this lower incidence of dengue in Ortigueira after the intervention with the NVC was statistically significant, we performed the measurement of monthly moving averages of dengue incidence in both Ortigueira (intervention area) and control cities. As can be seen in Figure 3B, the reduction in the number of dengue cases was reduced by up to 97.7% (CI −95.7 % to – 99.7 %) % over the intervention period, confirming the effectiveness of the method in preventing dengue.

## Discussion

In 2021 our group presented the NVC, a new vector control method based on SIT technology (16). The study was comprised of a controlled field trial, in which we presented the production method of NVC sterile male mosquitoes, as well as the safety and efficacy of the method in suppressing *Ae. aegypti* mosquitoes. In that study we performed two intervention periods (INT1 and INT2), in which the treatment and control areas were reversed between the INT1 and INT2 periods. In both intervention periods, the incidence of dengue was drastically reduced in the areas where the NVC was implemented, which showed that the NVC program is robust and not biased (16). Herein we extended the program to a large-scale intervention in an entire city in the State of Paraná, southern Brazil. Our objective now was reinforcing the effectiveness of the NVC method and to demonstrate the feasibility of a large-scale NVC deployment in real-life conditions.

Here we show a clear reduction in viable progeny of *Ae. aegypti* throughout the intervention period in Ortigueira (Figure 1). The substantial suppression of the mosquito population started about 15 weeks after the beginning of NVC intervention. The likely reason for the delay in the suppression of the field population is that the releases of NVC sterile mosquitoes started only at the end of November 2020 due to bureaucratic delays (authorizations and local regulatory approvals). This delay in suppression due to a late start (beginning of summer when mosquito populations already start growing exponentially), underscore the importance to begin the intervention as early as possible in the mosquito season, as has already been demonstrated in other SIT studies (9). In addition, our results indicate that an intervention period of at least 2-years is necessary for the full suppression effect to be achieved. This is probably due to the fact that in *Ae. aegypti* eggs may lie dormant for over a year (23). It is also important to note that during the period of intervention with NVC in Ortigueira, no other method of controlling the mosquito vector, such as the use of insecticides, was used. On the other hand, Tibagi requested to local authorities the use of Ultra-Low Volume insecticide spraying in public areas in an attempt to contain the spread of dengue in the city. Nevertheless, the incidence of dengue in the city was the worst among the control cities in the region (Figure 3A) (24).

Therefore, the reduction of the local population of mosquitoes and the consequent reduction of up to 150 times in the dengue incidence is exclusively due to the NVC intervention. This demonstrates the efficacy and facilitates ultra-low impact vector control and disease prevention in the face of climate change, while avoiding any collateral impact on biodiversity, which is the norm for any class of insecticides.

Another important aspect to consider after a large-scale NVC intervention is the possibility of mosquito reintroduction after suppression has been achieved. It is well known that the distribution of *Ae. aegypti* is largely driven by human movement in the presence of a suitable climate (25). Ortigueira is not an isolated city, and therefore there is daily movement of vehicles and people between Ortigueira and the neighboring cities. This represents the reality of the global economy, and therefore, to be effective, mosquito population suppression projects need to be performed on a large-scale. The solution to control the mosquito reintroduction in the area previously treated with NVC lies in maintaining entomological surveillance, particularly around high-risk introduction routes such as highways (25), and subsequently carrying out punctual releases of NVC sterile male mosquitoes in areas at greatest risk of mosquito reintroduction.

The mosquito suppression SIT methodologies, such as genetic modification to introduce a dominant lethal gene (9), cytoplasmatic incompatibility IIT (26,27) and more recently the NVC (16), have all shown reduction in the *Ae. aegypti* mosquito population. However, despite these numerous demonstrations of efficacy and safety, SIT is still not being used as a real-world strategy to combat the vector (28). One of the explanations for this gap between the development of the technology and its widespread implementation lies in the requirement to carry out at least 2 randomized controlled trials (RCTs) in two consecutive transmission seasons for products related to vector control, in order to provide sufficient evidence of the public health impact of these methods (1,29). Notwithstanding the theoretical strength of the data that may be generated in such RCT studies, they are expensive to carryout, time-consuming and most importantly - traditionally do not provide pragmatic evidence for how innovations are used or implemented in real-world practice (30). Therefore, financial, temporal and other resource restrictions may prevent such studies being conducted and stop valuable vector control tools from being rolled out (30). One possible solution for this gap was raised by Vontas and colleagues (2014) which hypothesized that once RCTs establish links between entomological and epidemiological indicators then rapid evaluation of new products within the same product category may be conducted through smaller scale experiments without repetition of lengthy and expensive RCTs (31). Currently, together with our emphatic results pertaining to the prevention of dengue, a plethora of additional evidence exists to support the immediate massive adoption and implementation of SIT technologies to meet WHO goals of dramatic reduction in the incidence of dengue.

The COVID-19 pandemic has highlighted several problems related to how health authorities address a crisis, whether related to emerging or existing diseases (32,33). Traditionally, new treatments, vaccines and prevention methods must go through several stages of testing and regulatory approvals, ranging from pre-clinical trials to commercial approvals before being widely used (34). In the pre-COVID-19 pandemic period, these processes could take more than 15 years to complete (34,35). The COVID-19 pandemic, however, has shown us that this process can be accelerated, and still prove safe and effective for the human population (36). The joining of forces between the different stakeholders involved in global health, which includes scientists, investors, regulatory authorities, and the industry promoted the de-bureaucratization of health and resulted in the relative containment of damage caused by COVID-19.

Dengue and other arboviruses have a massive global human and economic burden. In 2017, the WHO proposed a target of a 75% reduction in global dengue cases by 2027 (1). In the current trajectory, this goal will probably not be realized. On the contrary, an increase in the incidence of dengue has been observed over the last few years (37), and human induced warming phenomena such as flooding and heatwaves will likely further exasperate arboviral disease transmission (38). Therefore, new safe and effective vector control tools, like NVC, a SIT-based method, are urgently needed to reduce the burden of vector-borne diseases and mitigate threats to existing methods, such as ineffectiveness of insecticides (1). The suffering of hundreds of millions of people can and should be averted.

## Conclusions

Here we reinforced the effectiveness of NVC, in suppressing the *Ae. aegypti* mosquito population and protecting an entire city from a dengue outbreak that impacted the neighboring cities. Currently, the plethora of successful SIT interventions support their being implemented globally. We call on WHO and other stakeholders involved in prevention and treatment of arbovirus disease to immediately adopt existing technologies throughout the world to mitigate human suffering.

## Data Availability

All data produced in the present work are contained in the manuscript

## Notes

### Competing Interest Statement

All authors have completed the ICMJE uniform disclosure form at www.icmje.org/coi_disclosure.pdf and declare: DACF and CS has no interest conflicts, FAA, LCP, AR, DAO, BP, DR are employees of Forrest Brasil Tecnologia Ltda.; UP, MM, RAR, PB and ELR are employees of Klabin and NP is employee of Forrest Innovations Ltd.

### Clinical Trial

Environmental License Number 36127

### Funding Statement

This study was funded by Forrest Innovations and Klabin.

