## Supplementary figures for "Large-scale deployment of SIT-based technology in a Brazilian city prevented Dengue outbreak"

### Slide 1
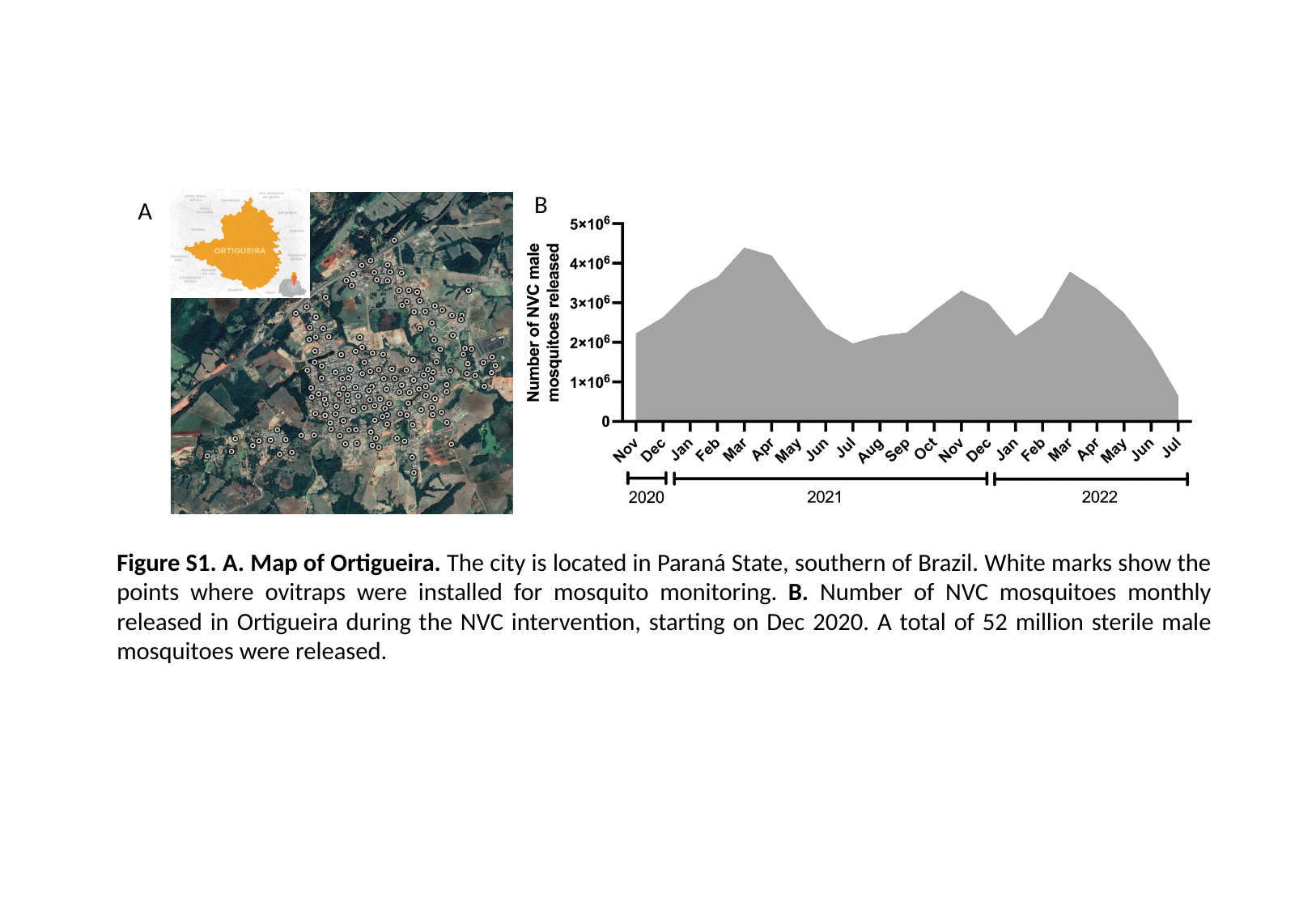

B
A
Figure S1. A. Map of Ortigueira. The city is located in Paraná State, southern of Brazil. White marks show the points where ovitraps were installed for mosquito monitoring. B. Number of NVC mosquitoes monthly released in Ortigueira during the NVC intervention, starting on Dec 2020. A total of 52 million sterile male mosquitoes were released.

### Slide 2
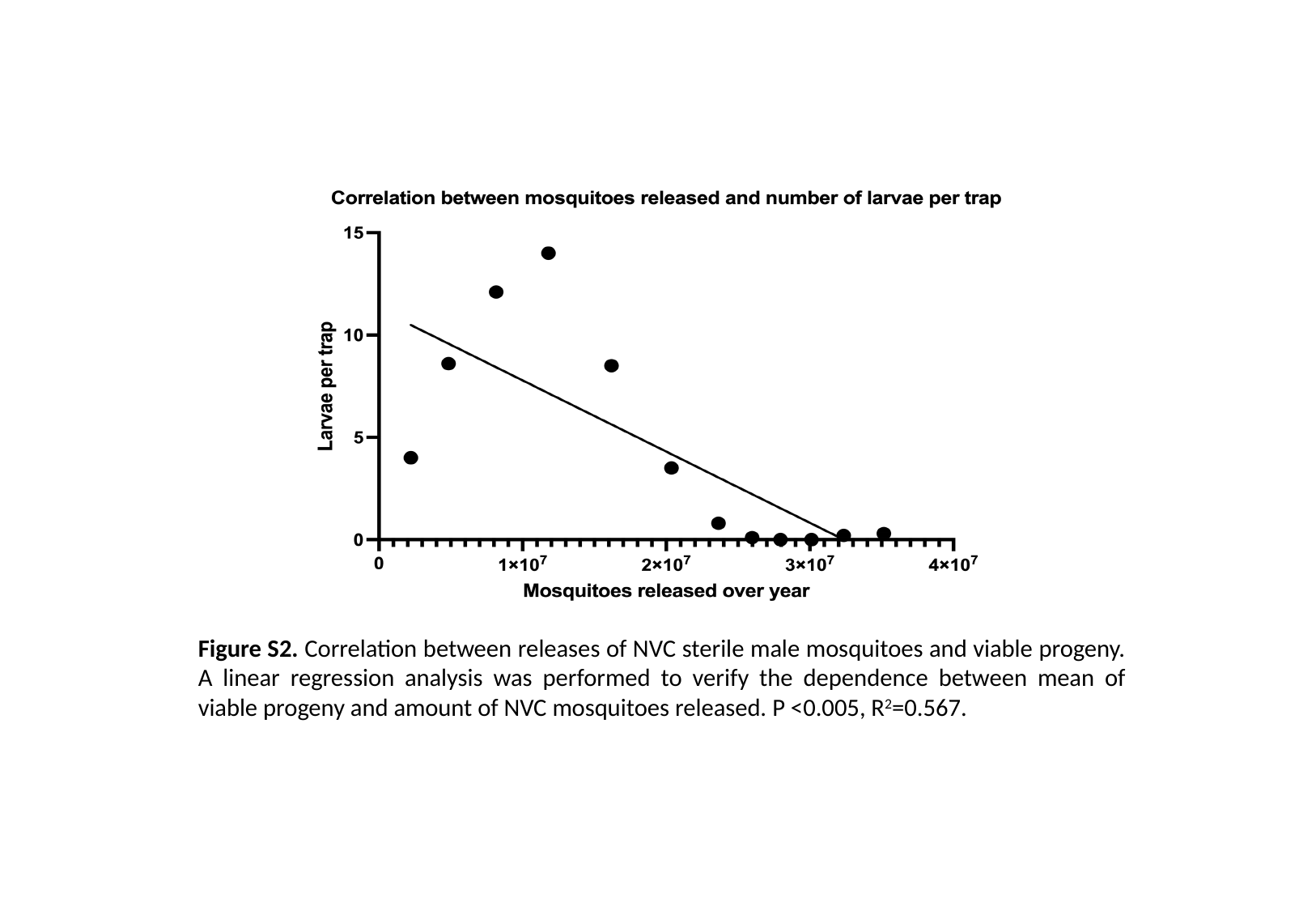

Figure S2. Correlation between releases of NVC sterile male mosquitoes and viable progeny. A linear regression analysis was performed to verify the dependence between mean of viable progeny and amount of NVC mosquitoes released. P <0.005, R^2^=0.567.

### Slide 3
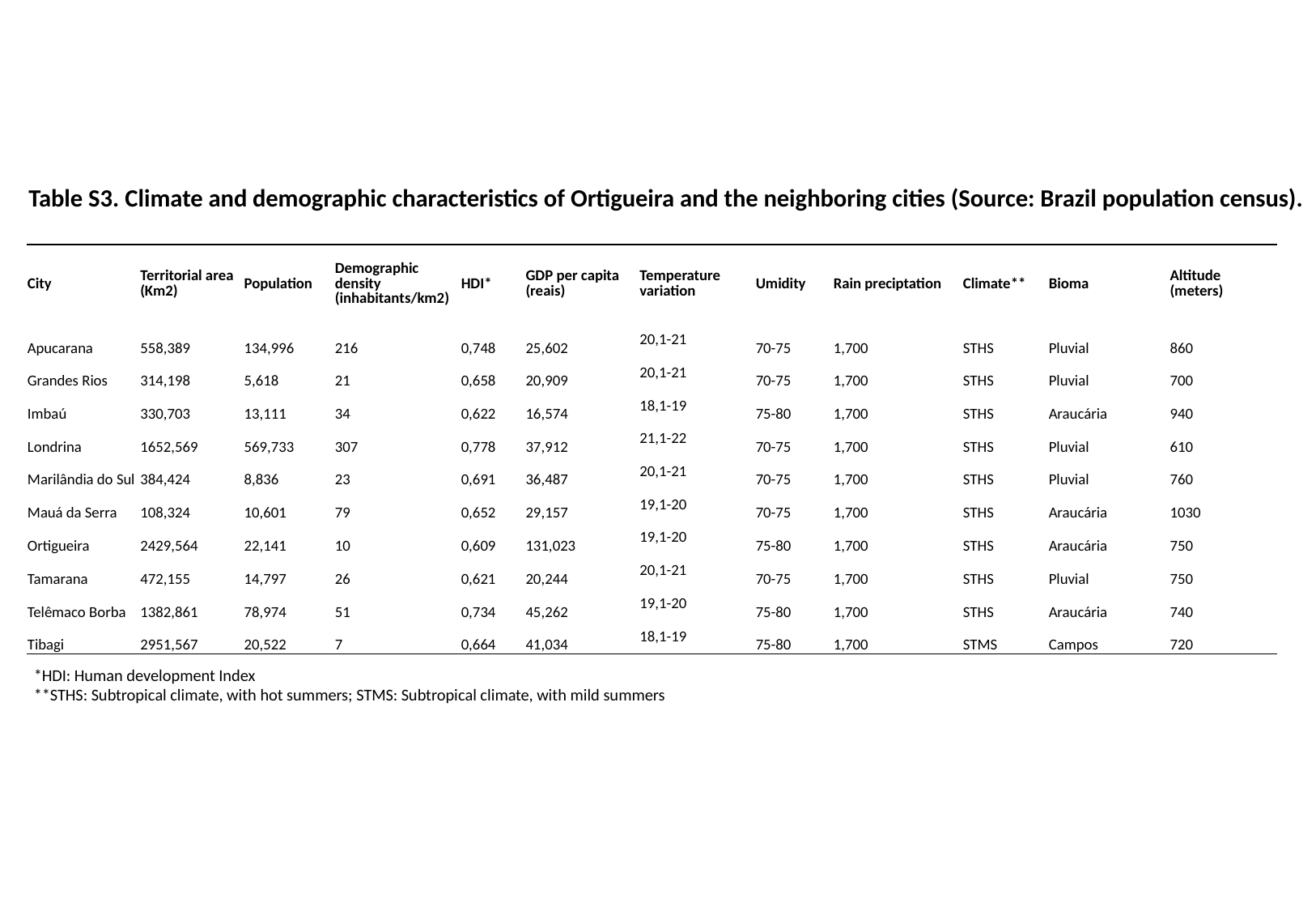

Table S3. Climate and demographic characteristics of Ortigueira and the neighboring cities (Source: Brazil population census).
| City | Territorial area (Km2) | Population | Demographic density (inhabitants/km2) | HDI\* | GDP per capita (reais) | Temperature variation | Umidity | Rain preciptation | Climate\*\* | Bioma | Altitude (meters) |
| --- | --- | --- | --- | --- | --- | --- | --- | --- | --- | --- | --- |
| Apucarana | 558,389 | 134,996 | 216 | 0,748 | 25,602 | 20,1-21 | 70-75 | 1,700 | STHS | Pluvial | 860 |
| Grandes Rios | 314,198 | 5,618 | 21 | 0,658 | 20,909 | 20,1-21 | 70-75 | 1,700 | STHS | Pluvial | 700 |
| Imbaú | 330,703 | 13,111 | 34 | 0,622 | 16,574 | 18,1-19 | 75-80 | 1,700 | STHS | Araucária | 940 |
| Londrina | 1652,569 | 569,733 | 307 | 0,778 | 37,912 | 21,1-22 | 70-75 | 1,700 | STHS | Pluvial | 610 |
| Marilândia do Sul | 384,424 | 8,836 | 23 | 0,691 | 36,487 | 20,1-21 | 70-75 | 1,700 | STHS | Pluvial | 760 |
| Mauá da Serra | 108,324 | 10,601 | 79 | 0,652 | 29,157 | 19,1-20 | 70-75 | 1,700 | STHS | Araucária | 1030 |
| Ortigueira | 2429,564 | 22,141 | 10 | 0,609 | 131,023 | 19,1-20 | 75-80 | 1,700 | STHS | Araucária | 750 |
| Tamarana | 472,155 | 14,797 | 26 | 0,621 | 20,244 | 20,1-21 | 70-75 | 1,700 | STHS | Pluvial | 750 |
| Telêmaco Borba | 1382,861 | 78,974 | 51 | 0,734 | 45,262 | 19,1-20 | 75-80 | 1,700 | STHS | Araucária | 740 |
| Tibagi | 2951,567 | 20,522 | 7 | 0,664 | 41,034 | 18,1-19 | 75-80 | 1,700 | STMS | Campos | 720 |
*HDI: Human development Index
**STHS: Subtropical climate, with hot summers; STMS: Subtropical climate, with mild summers

### Slide 4
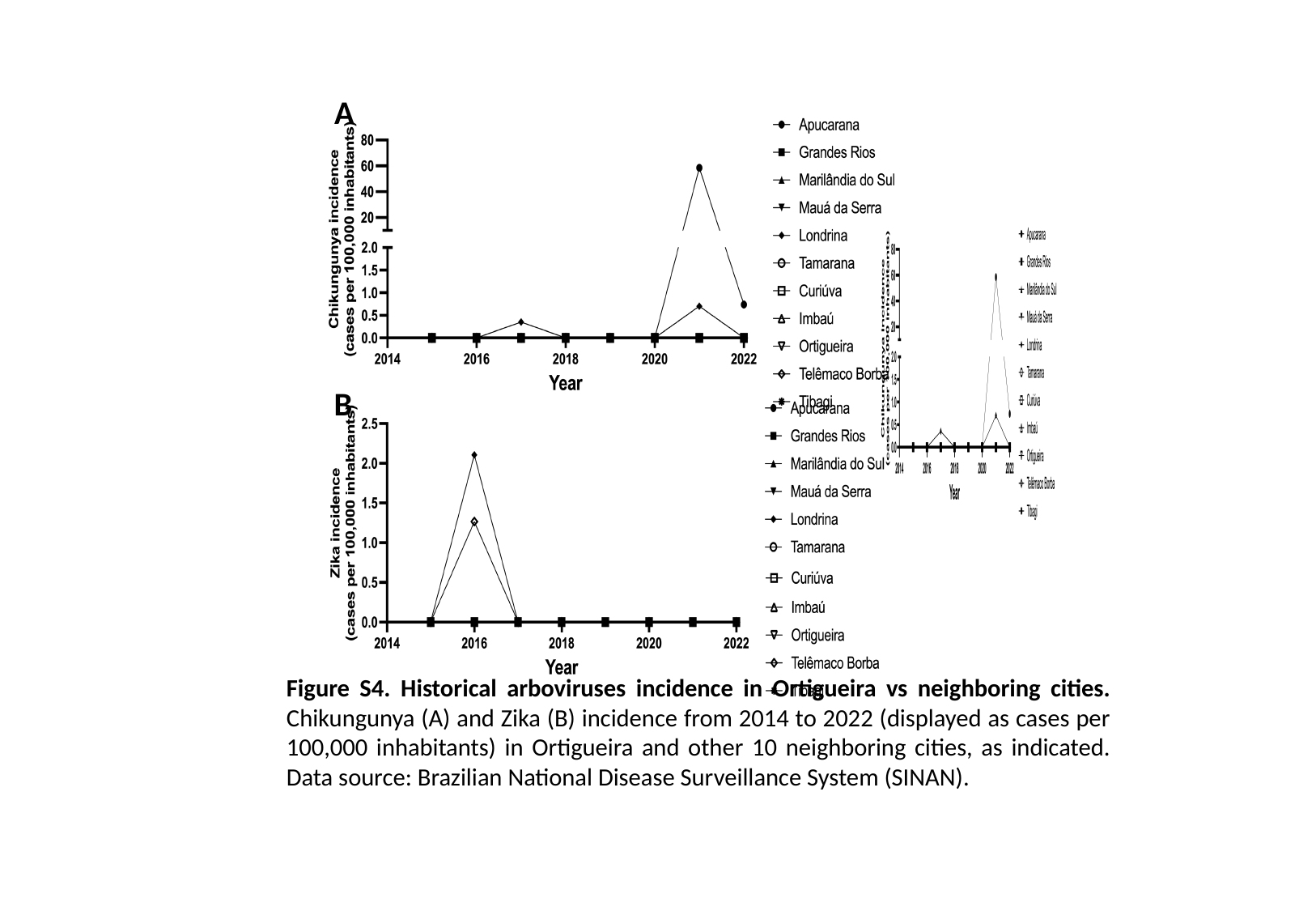

A
B
Figure S4. Historical arboviruses incidence in Ortigueira vs neighboring cities. Chikungunya (A) and Zika (B) incidence from 2014 to 2022 (displayed as cases per 100,000 inhabitants) in Ortigueira and other 10 neighboring cities, as indicated. Data source: Brazilian National Disease Surveillance System (SINAN).
